# Improving the impact of pharmacy interventions in hospitals

**DOI:** 10.1101/2023.03.03.23286757

**Authors:** Rafael Baptista, Mary Williams, Jayne Price

## Abstract

The interventions of pharmacy professionals are considered impactful inputs towards optimised patient care and safety, by rationalising prescriptions, enhancing therapeutic choices and reducing and preventing medication errors and adverse effects. Between October 2020 and October 2021, the community hospitals at Powys Teaching Health Board recorded 158 interventions, corresponding to 0.4 interventions per pharmacy staff per week. Predominantly, only two members of the team were recording these pharmacy interventions (PIs). Poor indicative PIs can result in lost opportunities for medication optimisation and prescribing rationalisation, increased costs and unidentified education and training potentials.

The aims of this project were (1) to record 180 interventions between 22/11/2021 and 08/04/2022 (20 weeks), corresponding to an average 3-fold increase, compared to the interventions recorded between October 2020 and October 2021 (52 weeks); (2) to have all hospital pharmacy staff recording at least one intervention during the same period.

The number of interventions recorded and the number of pharmacy staff recording each intervention were two process measures. The project was completed through two Plan-Do-Study-Act (PDSA) cycles and applied theory on managing change in healthcare.

The most successful intervention influencing positively the process measures was the implementation of a new <u>P</u>harmacy Intervention <u>R</u>ecord <u>T</u>ool (xPIRT) toolkit that included an online recording tool (xPIRT) and an interactive panel with up-to-date results from all interventions recorded (xPIRT Dashboard). Motivating change was proven to be one of the best determinants of user satisfaction and engagement that contributed to meet the project’s targets. xPIRT Dashboard provided staff the capacity to act on possible personal motivators and the possibility to improving care with medicines on their wards. The implementation of xPIRT toolkit was able to increase the representativity and significance of PIs recorded by the hospital pharmacy team and it is expected to be used for personal professional development, demonstrating team activity and impact, service planning, prescribing practice optimisation, and to identify education/training needs. This toolkit can be easily applied and adapted to other health organisations, settings and services.

## Problem and aims

This project focused on evidencing the impact of interventions made by pharmacy professionals (pharmacists and pharmacy technicians) in Powys Teaching Health Board community hospitals (PTHB), Wales. Optimising learning from these interventions will influence positively patient safety and quality of care.[1,2] Poor indicative pharmacy interventions (PIs) can result in reduced opportunities for medication optimisation and prescribing rationalisation, increased costs and unidentified education and training potentials.

Between October 2020 and October 2021, the Medicines Management (MM) team (MM) of PTHB have registered 158 interventions from two hospital pharmacy professionals — or 28.6% of the total number of the pharmacy team (PT) staff. This corresponded to an average of 0.4 interventions per pharmacist/pharmacy technician per week. These figures were not a true representation of the work that the hospital PT develop in Powys and failed to collect representative data.

The aims of this project were: (1) the hospital PT in Powys to record at least 180 interventions between 22/11/2021 and 08/04/2022 (20 weeks), corresponding to an average 3-fold increase of the number of PIs recorded between October 2020 and October 2021 (52 weeks); (2) to have all PT staff recording at least one intervention during the same period, corresponding to an increase of 75% in the number of staff recording interventions. These goals were based on staffing numbers, their working hours, and targets set up by the MM team in Powys.

## Background

The interventions of pharmacy professionals are considered valuable inputs towards optimised patient care and safety, by rationalising prescriptions and reducing and preventing medication errors.[3] Multiple reports have concluded that pharmacy professionals were able to reduce the length of stay of hospital inpatients[4] reduce readmission rates,[4,5] decrease the number of adverse drug reactions,[6] promote better medicines use and adherence,[7–9] and discontinue inappropriate prescribed medicines.[10]

Besides the consensus on the positive clinical impact of PTs, more cost-consequence analyses are needed to evaluate the economic outcomes of these interventions. Nonetheless, several research articles and meta-analysis concluded PIs contribute for more cost-effective prescribing choices across a range of clinical conditions.[11–14] The economic impact of PIs may inform stakeholders and decision-makers for policies that support more pharmacy funding to hospitals and community pharmacies.[15]

Several strategies have been used to assess the clinical impact of PTs worldwide. The All Wales Intervention Database (AWID) is available for the health boards in Wales. The Pharmacists Achieve Results with Medications Documentation (PhARMD), a template that captures interventions and outcomes of clinical pharmacists in an integrated healthcare system, is currently being used in several hospitals in the United Stated of America.[3] Other recording tools have been previously reported.[16–18]

## Baseline measurement

An initial baseline study was conducted on the recorded interventions by the hospital PT in Powys between October 2020 and October 2021. The only available tool used to collect this information was AWID. Data collected — date, number of interventions and identification of the contributor — was organised in a Microsoft Excel spreadsheet. A total of 158 interventions was recorded by two pharmacy professionals. The calculated median was zero due to the lack of consistent data throughout the period (Figure 1).

**Figure 1.**
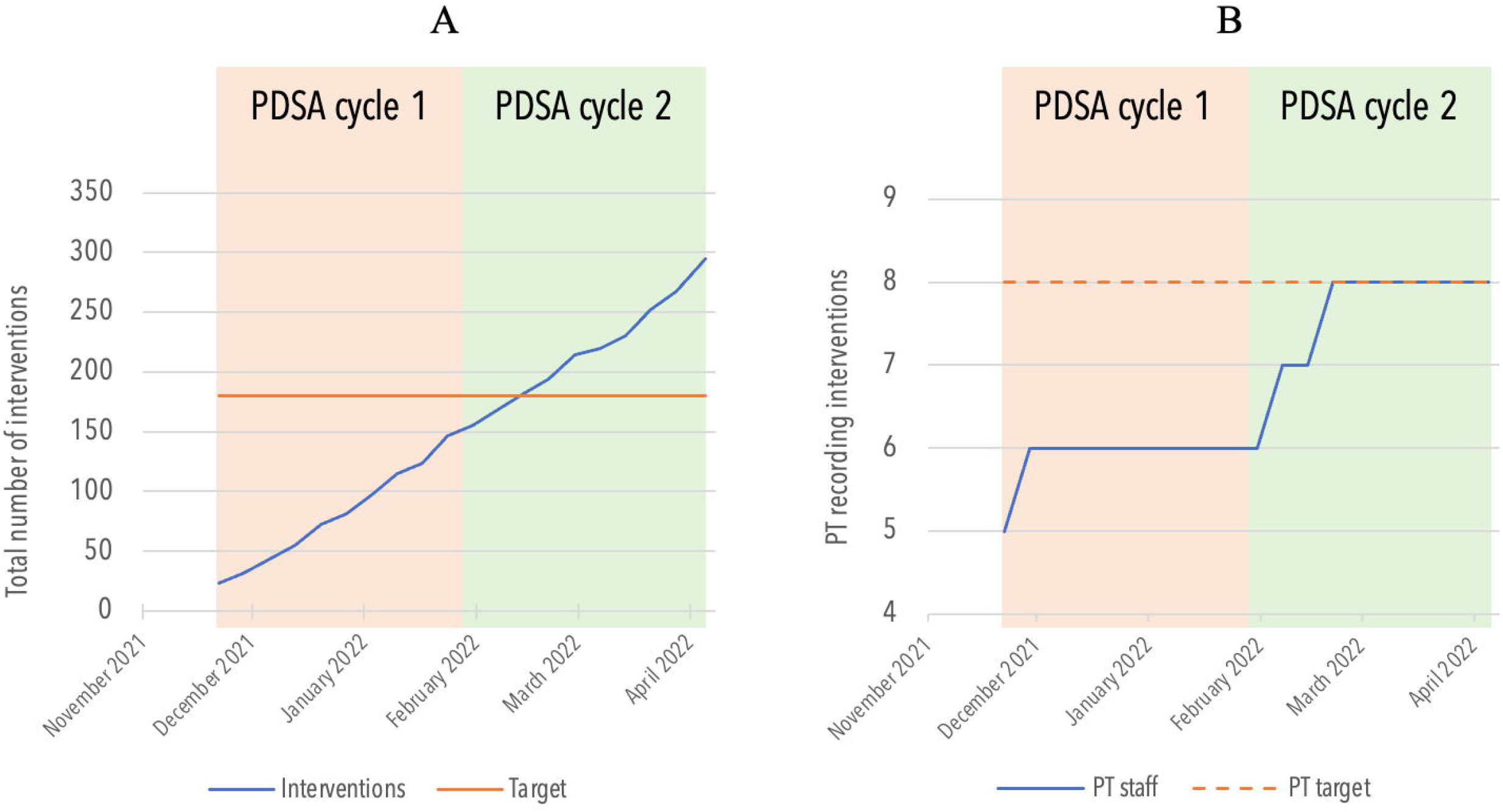
A - Total number of interventions recorded on xPIRT (blue line) and target established for the project (orange line). The target was met during PDSA cycle 2. B – Total number of pharmacy team (PT) staffing recording interactions via xPIRT (blue line) and target established for the project (dashed orange line). This target was met during PDSA cycle 2.

## Design

The first author of this report was secured as ‘champion’ for this project and implemented all PDSA methodology and quality improvement strategies. The PIs recorded by the author were not considered in the project, reducing the risk of bias.

The process measures of the project were the number of interventions recorded and the number of staff recording them. The outcome measures were the number of new local guidelines being considered/written for Powys’ hospitals, based on the interventions recorded, and the awareness level of the PT for recording PIs.

In this project, it was important to understand unintended consequences of the changes that were implemented. Thus, the average time taken to record one intervention was indicated as a balancing measure, to ensure that recording interventions would not have a considerable impact in the existing pharmacy services on the wards of Powys hospitals. The time taken to record interventions was automatically retrieved via Microsoft Forms.

## Strategy

Two Plan-Do-Study-Act (PDSA) cycles were undertaken over a 20-week period. Table 1 provides a summary of these details.

**Table 1.** Overview of the study design with specific changes in each PDSA cycle listed.

| PDSA cycle | Contributors for inadequate record of PI | Changes |
| --- | --- | --- |
| PDSA 1<br>(10 weeks) | Existing tools – AWID – are not easy to use. | Implementation of a new <u>Pharmacy Intervention Record Tool</u> — xPIRT — via Microsoft Forms. |
|  | MM leadership did not inform the team of the importance of PI. | To remind and motivate the PT every Monday, via email. |
|  |  | Any small changes on xPIRT should be implemented within 24 hours, after emailed feedback. |
|  | Time limitation. | Reassurance that xPIRT is an easy-to-use form that would take less than 5 minutes to complete per intervention. |
| PDSA 2<br>(10 weeks) | Interventions' results were not accessible for users. | Implementation of xPIRT Dashboard (via Microsoft Power BI), where all instantly updated results are readily available to all PT, via Microsoft Teams. |
|  | Intervention's results were not editable. | Creation of a sharable file, xPIRT List, (via Microsoft Lists) where results can be editable. |
|  | Lack of awareness on the purpose/use of the data from interventions. | PT advisory meeting with intervention of the Head of Community Services/Pharmacy about the use of xPIRT and its data – local clinical governance meetings, inform local clinical guidelines and training needs. |
|  | Time limitation. | PT advisory meeting with reassurance that, in order to meet the project's targets, each PT staff would only need to invest 3 minutes per week on xPIRT. |

AWID – All Wales Intervention Database; MM – Medicines Management; PDSA – Plan-Do-Study-Act; PT – pharmacy team; xPIRT – Pharmacy Intervention Record Tool.

## Plan-Do-Study-Act (PDSA) cycle 1

To complement the baseline data, two surveys were completed by stakeholders (PT members, ward managers and ward doctors), to evaluate the understanding and satisfaction with the current methods of recording PIs.

All ward managers and prescribers initially surveyed agreed or strongly agreed that pharmacy interventions change practice, improve patient safety and support service planning, and would make them more aware of the role of the pharmacy teams in hospitals.

A stakeholder analysis was completed to establish priorities and differentiate stakeholders based on their power/ interest in the project. Although essential stakeholders, ward managers and prescribers were considered to have low power in the project, as this change focused more on the pharmacy teams. In order to meet the aims of the project, these stakeholders would only be consulted in the beginning of the project.

The PT stakeholders revealed that 75% of the hospital PT have not recorded their interventions in a platform that maintained data security, between October 2020 and October 2021. Half of the PT did not record any interventions at all. A fishbone diagram (Supplementary Figure 1) depicted the issues encountered with the low number of interventions recorded. Further analysis showed that time limitation, poor existing recording tools – AWID –, and lack of information or targets set up by the MM leadership were the key factors that influenced the inadequate number of documented PIs (Supplementary Figure 2).

Following the collection of the baseline data, our findings were presented to the PT stakeholders, to encourage feedback and suggestions for improvement. Using the baseline data and feedback received in the meeting, a PDSA methodology was implemented to complete a quality improvement strategy over a 20-week period. Each PDSA cycle included a survey completed by all PT following by a short planning meeting for loop feedback.

A short meeting and feedback loop, with baseline data for PIs recorded between October 2020 and October 2021, was held with PT stakeholders in the beginning of PDSA cycle 1. After suggested changes were in place, data was collected using the new <u>P</u>harmacy Intervention <u>R</u>ecord <u>T</u>ool — xPIRT (Supplementary Figure 3) — for 10 weeks, from 22/11/2021 to 28/01/2022. xPIRT was developed using Microsoft Forms. Punctual feedback and additional suggestions were sent via email and resolved within the same day.

## PDSA cycle 2

A short meeting and feedback of an advisory survey took place before recording interventions for the second PDSA cycle. A driver diagram was built to better visualise the changes that needed to be implemented (Supplementary Figure 4).

In addition to the use of xPIRT, started in PDSA cycle 1, changes discussed in the meeting included the assessing and editing of intervention results. Thus, an editable file available online via xPIRT List with real-time recordings was shared with the PT, along with xPIRT Dashboard (Supplementary Figure 5), available on Microsoft PowerBI (Premium License). Thus, xPIRT Toolkit was then formed by xPIRT, xPIRT List and xPIRT Dashboard. This enabled the team to keep track of all interventions done and select them, interactively, by ward, contributor, date, drug, and severity. The overall intervention recording pathway is depicted in Supplementary Figure 5.

Other topics were addressed in the short meeting. Due to the reported time limitations, it was reassured that, in order to meet the goals of this project, all PT staff should only spend 3 minutes per week recording interventions, based on the Microsoft Forms calculated response time per submitted record. Also, due to questions related with accuracy of the data recorded, the project ‘champion’ clarified his interventions were not considered when measuring improvement. Finally, due to feedback obtained, it was explained that this data collection would be valuable to use in local clinical governance meetings, would inform specific local guideline needs and could be used for personal continuing professional development (CPD).

After all suggested changes were put in place, data was collected using xPIRT for 10 weeks (from 31/01/2021 to 08/04/2022). A final survey was sent to the PT stakeholders to evaluate and feedback on the changes implemented during the project.

## Results and Discussion

In the first PDSA cycle, an increase in the average number of interventions recorded (14.7 interventions per week) was registered, 63.3% above the target set for end of the project. The median number of interventions recorded rose to 12, indicating a clear improvement from the baseline. However, during this cycle not all the hospital PT, only 6 out of 8, have used xPIRT. Nonetheless, this cycle provided valuable information that the changes impacted positively on the number of interventions completed (Figures 1 and 2).

**Figure 2.**
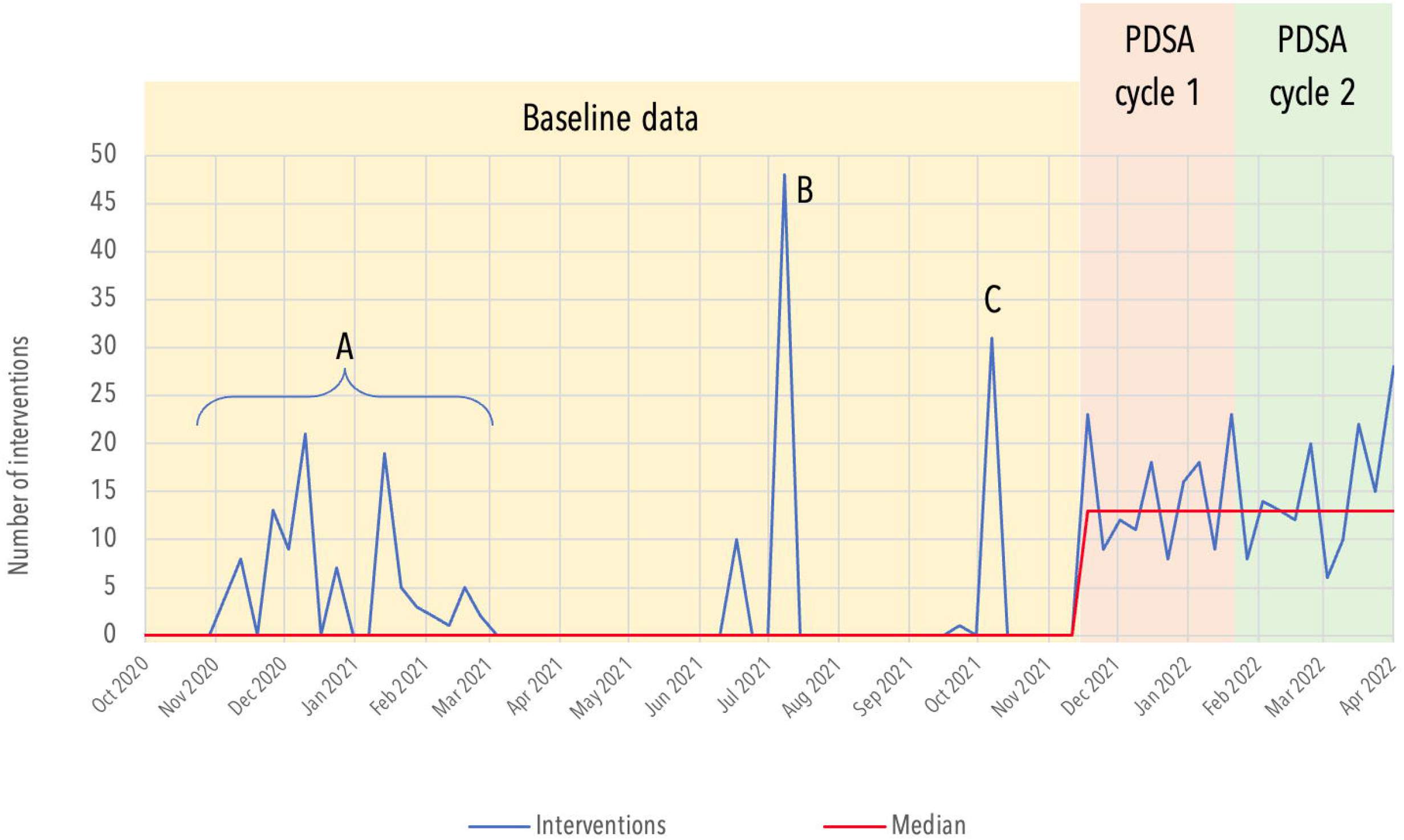
Number of interventions recorded on xPIRT and respective median. Background data was obtained via All Wales Intervention Database (AWID): interventions recorded in ‘A’ represent an attempt to use AWID, showing to be unsuccessful after March 2021; astronomical points ‘B’ and ‘C’ demonstrate that AWID does not allow the user to choose the interventions dates, thus concentrating all recordings in the same date, when interventions are submitted. PDSA cycle 1 and 2 are evidenced with the red and green areas, respectively.

The second PDSA cycle also saw an increase in the average number of interventions recorded (14.8 interventions per week), 63.9% above the target set for end of the project. The median number of interventions remained 12, indicating a clear improvement from the baseline but no different from PDSA cycle 1. A total of 254 interventions were recorded, 41.4% above the target. This target was achieved seven weeks before the end of the project (Figures 1 and 2).

Both aims of the project were met six weeks in advance of the end of the second PDSA cycle, signified when all members of the PT had recorded at least one intervention (Figures 1 and 2).

Act-IP©, a website where French hospital pharmacists record their interventions,[19] was developed in 2006, due to the lack of validated tools for documenting PIs in hospitals in France. This website has recorded more than 800,000 PIs across 1100 hospitals by 3800 pharmacists, over the last 16 years. This enabled the study of key issues in pharmacy practice such as identifying trends in interventions on groups of medicines, diseases or medical speciality, and determining factors associated with prescribers’ PI acceptance.[20,21] As the authors concluded that most of the available tools did not measure the clinical, economic and organisational impact of PIs, the CLEO (Clinical, Economic and Organisational) scale was adapted Act-IP© v2.[18] However, the authors reference that the standardisation of documentation systems (e.g. information technology tools) of most of the PI recording tools are still defective.[22] With our project, the xPIRT Toolkit allowed integration of an easy and accessible tool which records clinical data and translated it to indicators of economic and organisational impact. The creation of the xPIRT Toolkit considered not only the identified needs for PI recording tools but was also driven by theory on managing change in healthcare.

By adopting a ‘systems thinking’ approach, people, tasks, technology, environment (physical, cultural, social) and organisational structures were all considered in the project.[23] The following considerations, explained below, were taken into account: (1) foundation concept, (2) multiple perspectives, (3) work conditions, (4) decisions’ explanations and understanding and (5) analysis, interactions and performance variability and flow.

A clear understanding of the concept of the project by stakeholders with power was essential for a positive impact in quality. Improving knowledge dissemination contributes not only for better clinical practice but also for enhanced performance in service improvements.[24] Knowledge translation in this project was achieved by making explicit the purpose and structure of the communication methods.[24] This project used a range of active-passive interactions to convey information: infographics, workshops, emails and short talks. All different strategies allowed the PT stakeholders to understand the importance of recording PIs, an important outcome measure. From the baseline data gathered, 87.5% of the PT suggested that the record of PIs was important, with this figure rising to 100% at PDSA cycle 2. Additionally, the same group of stakeholders considered that the advantages of recording PIs would outweigh possible disadvantages (from a baseline of 49.9% of the PT, to 85.7% at PDSA cycle 2).

Multiple perspectives from all stakeholders were considered. The use of anonymous staff feedback and rapid engaging techniques are known to contribute for improved service quality.[25,26] The use of anonymous surveys, in PDSA cycles 1 and 2, and continuous engagement via weekly emails, allowed the project to take on board essential feedback that contributed for building the xPIRT toolkit. In the short talk post-survey analysis, all ideas were considered but only some were applied. However, no discarded ideas were left without an adequate explanation. At the PDSA cycle 1, 80% of the PT considered that all their suggestions were considered, with this figure rising to 85.7% by the second PDSA cycle.

A focus was also given to the PT work conditions. The average time to complete one intervention, an important balancing measure of the project, was calculated to be in average 2.46 min in PDSA cycle 1 and 2.42 min in PDSA cycle 2. Although the average time to record one intervention did not change significantly between PDSA cycles (*P* = 0.903, t=0.1221, df=285, independent unpaired t-test), 83.3% of the PT indicated that recording PIs were a challenging competing priority with ward work at PDSA cycle 1, with this figure decreasing to 71.4% at PDSA cycle 2. This was achieved by avoiding the interesting concept of ‘projectness’ referred by Dixon-Woods et al.[27] Exhaustive project-centric ideas or conversations were avoided during surveys or short talks, reducing the risk of alienating staff. By acknowledging that PI would take some time from ward work and by keeping goals realistic and achievable, the PT staff was able to successfully participate and engage with the project.

The last point of ‘systems thinking’ regarding analysis, interactions and performance variability and flow was considered by applying three effective approaches: (1) relationship-, (2) motivational- and (3) value-based.

As relationship-based quality improvement strategies are often well succeeded due to easier coordination processes[28] this project made sure that tight communication channels were kept with every single PT staff.

The theory of motivating change describes the psychosocial-structural conditions for sustained change from the perspective of front-line staff. A total of 85.7% of the PT staff mentioned that xPIRT Dashboard would motivate them to record more interventions. The project intended to narrow the gap between intrinsic and extrinsic motivators. xPIRT Dashboard provided staff the capacity to act on possible personal motivators and the possibility of improving care on their wards. In addition, witnessing effective change was motivating for the staff as positive outcomes provided a convincing argument for the need to sustain improvement activity [29]. For instance, xPIRT Dashboard allowed the identification of four different potential needs for local guidelines or formulary changes. This has been identified as another important outcome measure. Motivating change along with the implementation of xPIRT Dashboard may justify the success of the project in meeting both targets during the second PDSA cycle.

Lastly, a value-based approach was linked to the ‘systems thinking’ as it empowers frontline teams and can achieve a transformational outcome.[30] xPIRT Dashboard allowed each member of the PT to consult cost avoidance projections by recording their interventions, per member of staff and as a group. This not only improved extrinsic motivation but also improved team culture and project engagement, towards a continuous quality improvement.

Overall, the PT satisfaction on this project, an important outcome measure, improved from the baseline of 2.5/5 to 4.9/5 stars, at PDSA cycle 2.

## Limitations

The significant variability within the PT presented as an important limitation for accurately assessing improvement, in this project. These variables included staffing numbers and profile, the number of days per week with ward cover by PT, and the information technology (IT) skill-mix.

It is known that recording interventions would have an impact on the PT ward work, as this could draw staff’s attention from patients. Although this was constantly fed back throughout the project, it is yet to be known the impact of this expected drawback.

This project was based on the work of the PT in community hospitals Powys and have not yet been transposed to primary care or district general hospitals. Also, the technology behind xPIRT Toolkit are dependent on Office 365 and Microsoft Power BI Premium licensing, which may be a limiting step for some pharmacy teams.

While it is known that PIs contribute for improved patient care and safety, as they optimise prescriptions and reduce and prevent medication errors, this measure of patient care and safety was not directly assessed throughout this project. A higher number of PI yields more representative data, true data, and xPIRT Toolkit can identify safety risks or unsafe care. However, it does not estimate harm, disability or death due to adverse events in inpatients.

## Conclusions

In the beginning of the PDSA cycle 1, it was highlighted that the record of PI lacked representativity and significance. Besides the low number of interventions recorded, the team was not motivated and did not understand the importance of recording this data.

The results here presented showed that an effective recording tool, that reflects the organisation’s wants and needs, is key for increasing the number of PI documented. xPIRT Dashboard created an automatically updated visual platform that organised data into infographics ready to be used for personal CPD, evidencing activity and impact, allowing service planning and informing clinical governance meetings, and identifying education/training needs.

Although it was reported that recording interventions may have drawn staff’s attention from patients, it is yet to be known the impact of this expected drawback. Future work will evaluate the additional workload potentially associated with the use of xPIRT Toolkit. The long-term success of this project after PDSA cycle 2 will continue to be monitored.

This toolkit can be easily applied and adapted to other health organisations, settings and services and is expected to contribute positively to patient safety.

## Supporting information

Squire Checklist

## Data Availability

All data produced in the present study are available upon reasonable request to the authors.

## Acknowledgments

The authors would like to thank all hospital staff of the medicines management/pharmacy, Powys Teaching Health Board.

## Contributors

Not applicable.

## Ethical approval

Not applicable.

