## Supporting Information for "Improving the impact of pharmacy interventions in hospitals"

Supplementary Information

**Supplementary Figure 1 –** Fishbourne diagram depicting issues encountered with the low number of PI recorded.

Low record of pharmacy interventions

Staff

Lack of understanding on the advantages of recording interventions

Time limitation

No protected time to complete this task

System/materials

AWID does not accept retrospective data

AWID is not easy to use

AWID is “clunky” and slow

AWID is not adapted to Powys hospitals

Management

Did not inform other team members about recording interventions

Did not motivate hospital teams to record interventions

Targets for PI have not been defined

AWID – All Wales Intervention Database; PI – Pharmacy interventions.

**Supplementary Figure 2-** Pareto chart highlighting frequency of key contributors for inadequate or low record number of pharmacy interventions.

Number of respondents

Cumulative percentage

|  | Count | Cumulat% |
| --- | --- | --- |
| Time limitation | 6 | 37.5 |
| Existing tools are not easy to use | 4 | 62.5 |
| MM leadership did not inform me | 3 | 81.25 |
| MM leadership did not motivate me | 2 | 93.75 |
| I do not understand the advantages of recording interventions | 1 | 100 |
| There is no tool to record my interventions | 0 | 100 |
| I am not motivated | 0 | 100 |

MM – medicines management.

**Supplementary Figure 3 –** **P**harmacy **I**ntervention **R**ecord **T**ool — x**PIRT** main page.

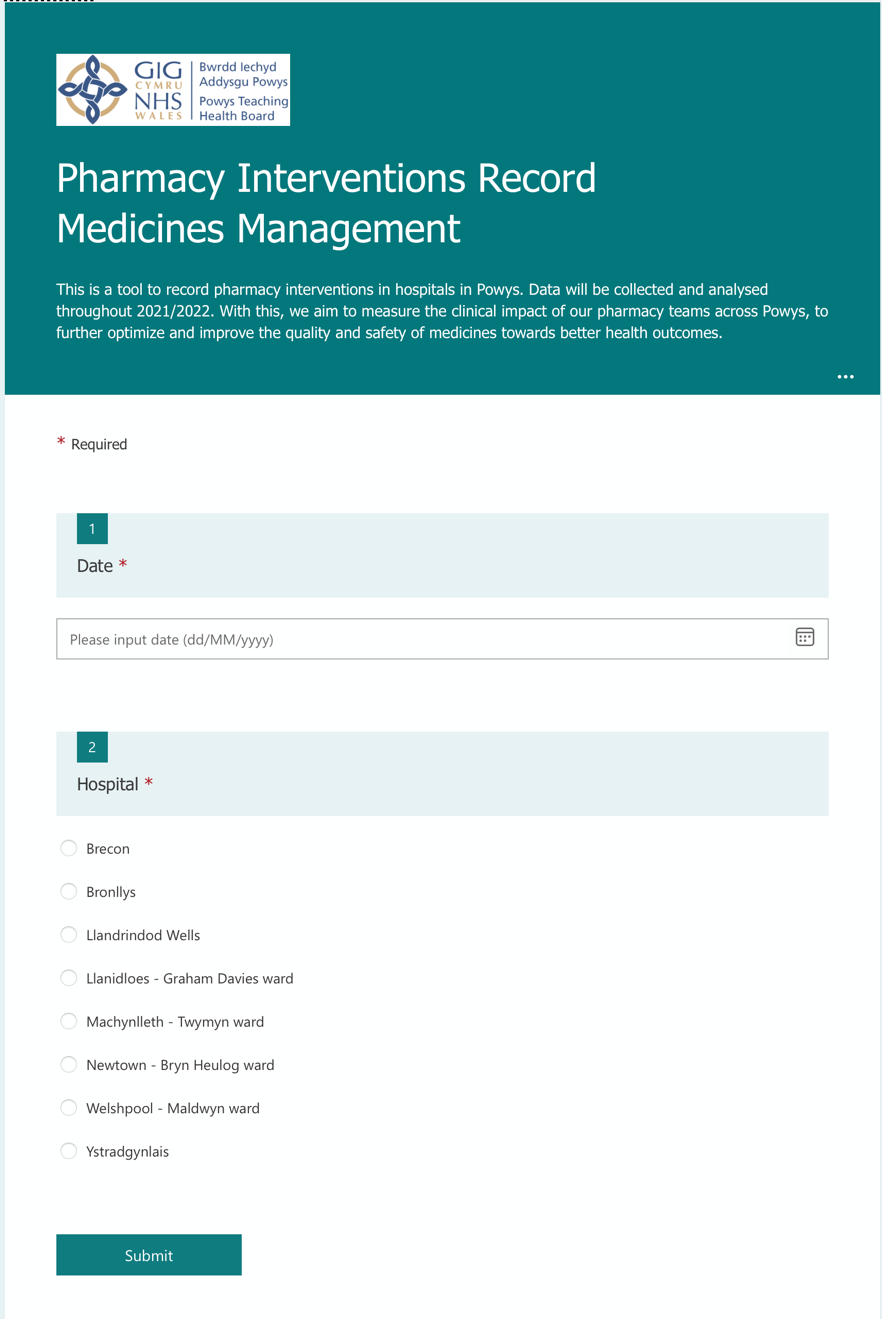

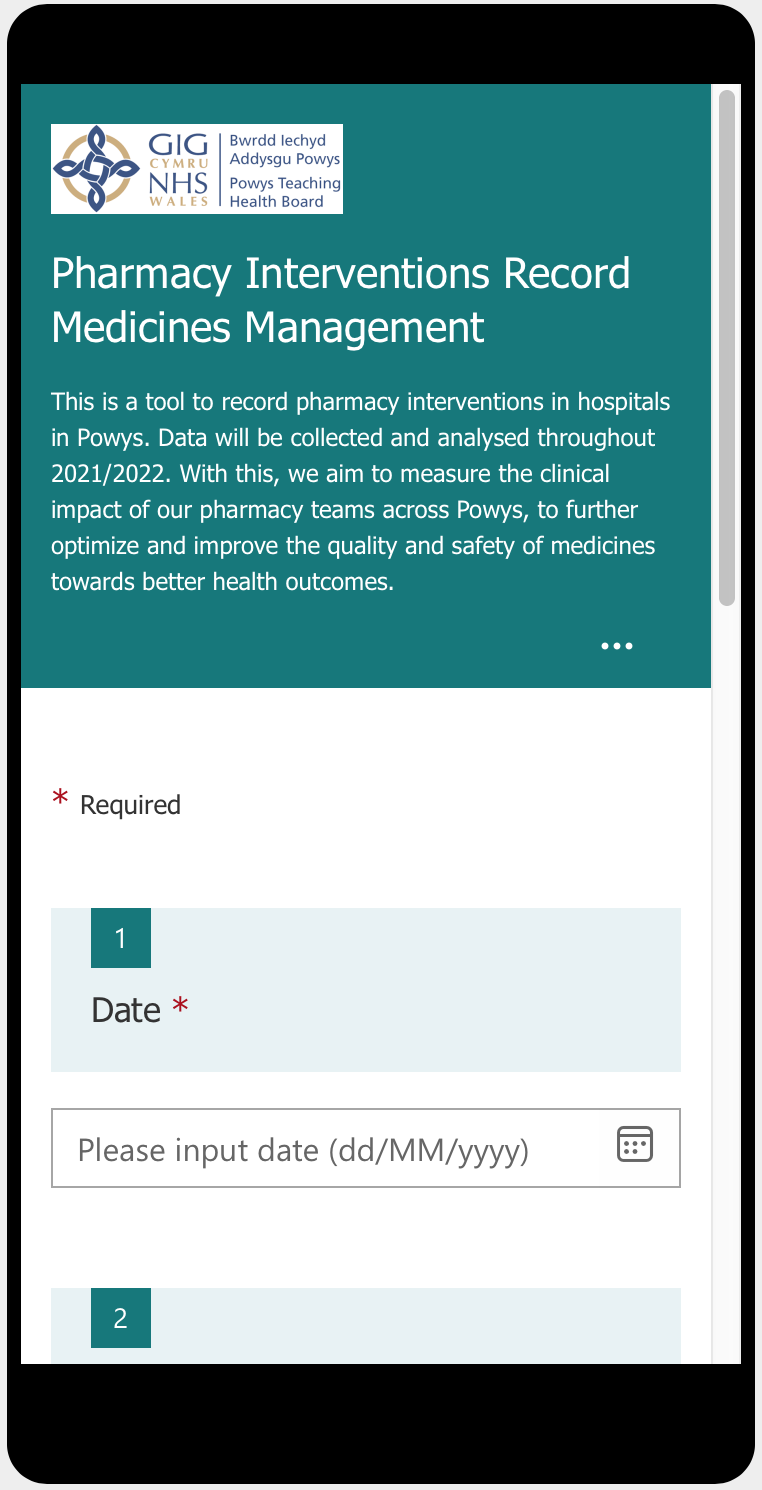

**Supplementary Figure 4 -** Driver diagram depicting changes to be implemented in PDSA cycle 2.

To record 180 interventions until 08/04/2022.

AND

To have all pharmacy team staff recording at least one intervention until 08/04/2022.

Recording PI

Easy to use recording tool

Leadership support

Interventions results to be easily accessed

Interventions results to be easily editable

Tackle time limitations

xPIRT can be used on a computer or phone

xPIRT Interactive Board in place

To share an editable file with interventions’ results

To address in the meeting: only required 3 min per week per staff, to meet goal

Weekly reminder email to be sent

To address in the meeting: data to be used in clinical governance meetings, to inform the need for specific local guidelines and for CPD.

To address in the meeting: PI made by project’s ‘champion’ are not considered for the project

Team to be reminded of recording interventions

Set clear goals regarding data usage

Clarify project ‘champion’’s bias in measuring improvement

PI – Pharmacy interventions; CPD – continuing professional development.

**Supplementary Figure 5 –** xPIRT Dashboard.

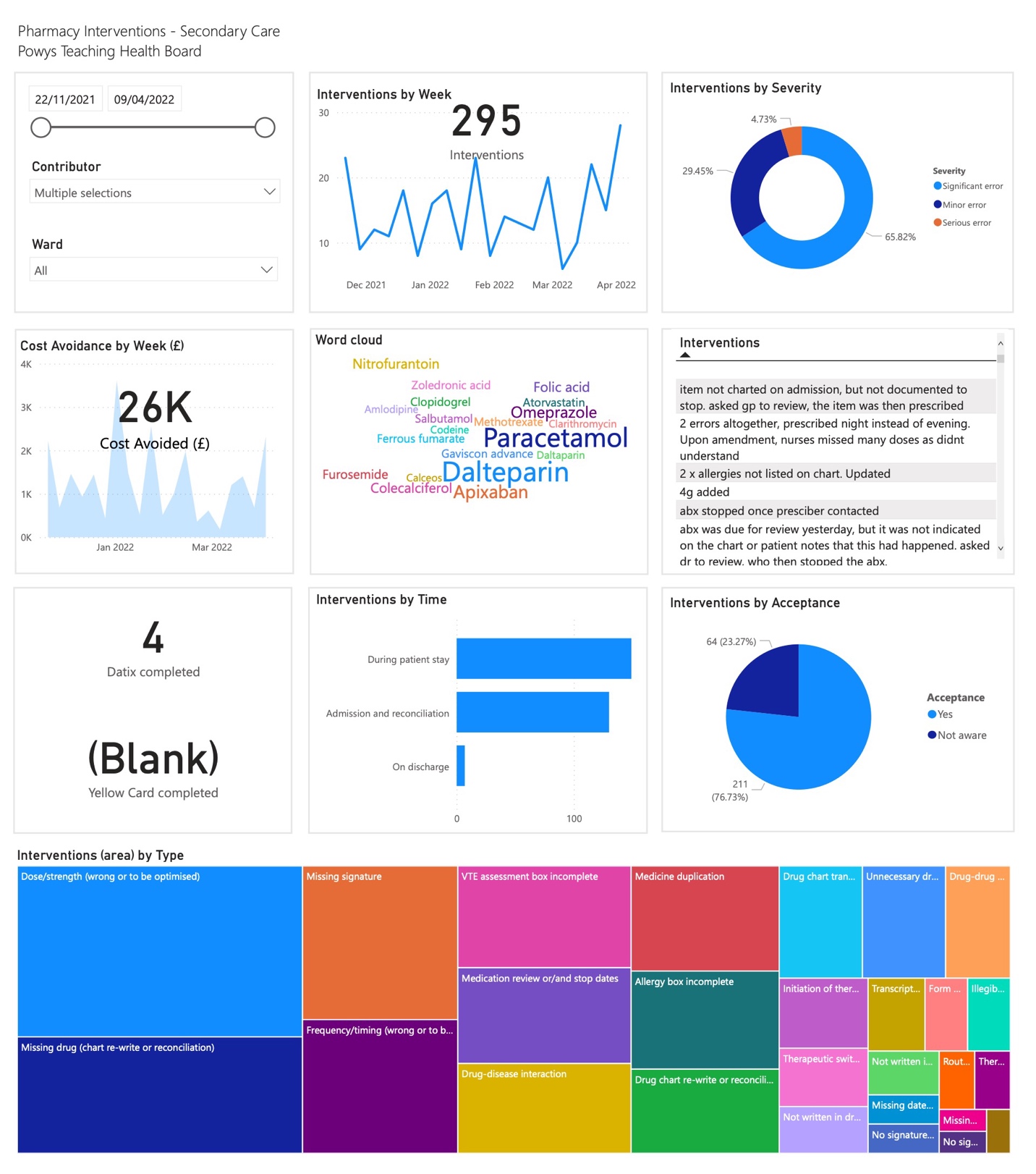

**Supplementary Figure 6 –** Flow chart with process pathway following a PI.

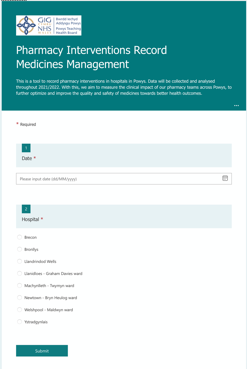

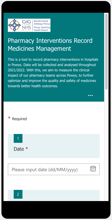

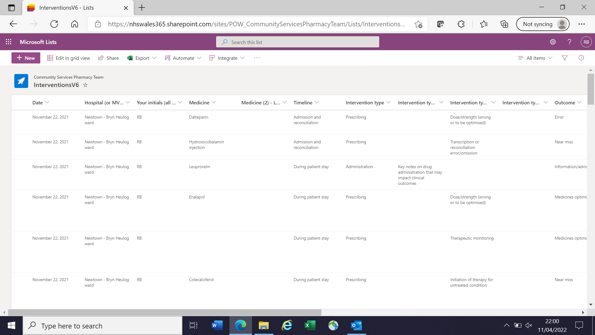

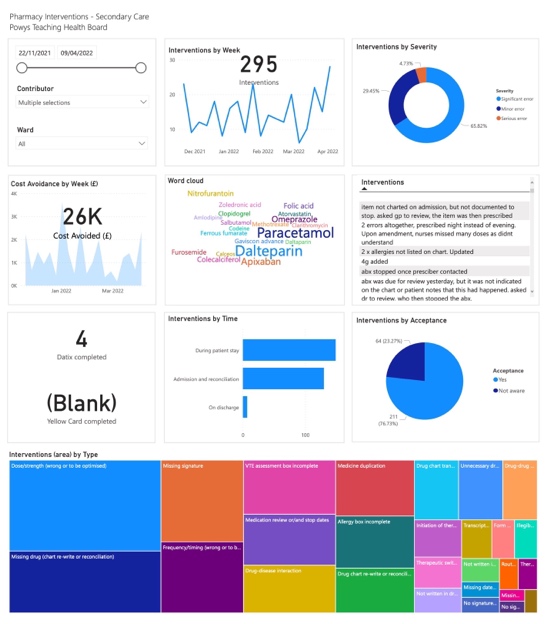

Recording tool

Average recording time is 2 minutes.

Data is organised in an automatic table that can be editable anytime.

Interactive Dashboard in available to our team and is updated every hour, based on xPIRT List.

This can be shared with ward and prescribers.

xPIRT

xPIRT List

xPIRT.Dashboard

xPIRT Toolkit
