## Supplementary material for "Improving the impact of pharmacy interventions in hospitals": Squire Checklist

| **Checklist SQUIRE 2.0** |  | |
| --- | --- | --- |
| **Text section and item name** | | **Page/line no(s).** |
|  | | **info is located** |
| **Title and abstract** | |  |
| 1. **Title** | |  |
| Indicate that the manuscript concerns an initiative to improve healthcare (broadly defined to include the quality, safety, effectiveness, patient-centredness, timeliness, cost, efficiency and equity of healthcare). | | 1 |
| 2. **Abstract** | | 1 |
| a. Provide adequate information to aid in searching and indexing. | | 1 |
| b. Summarise all key information from various sections of the text using the abstract format of the intended publication or a structured summary such as: background, local problem, methods, interventions, results, conclusions. | | 1 |
| **Introduction: Why did you start?** | | **2** |
| 3. **Problem description** - Nature and significance of the local problem. | |  |
| 4. **Available knowledge** - Summary of what is currently known about the problem, including relevant previous studies. | | 2 |
| 5. **Rationale** - Informal or formal frameworks, models, concepts and/or theories used to explain the problem, any reasons or assumptions that were used to develop the intervention(s) and reasons why the intervention(s) was expected to work | | 2 |
| 6. **Specific aims** - Purpose of the project and of this report. | | 2 |
| **Methods: What did you do?** | | 2-7 |
| 7. **Context** - Contextual elements considered important at the outset of introducing the intervention(s). | | 2 |

| 8. **Intervention(s)** | 5-7 |
| --- | --- |
| a. Description of the intervention(s) in sufficient detail that others could reproduce it. | 2-7 |
| b. Specifics of the team involved in the work. | 2-7 |
| 9. **Study of the intervention(s)** | 2-7 |
| a. Approach chosen for assessing the impact of the intervention(s). | 2-7 |
| b. Approach used to establish whether the observed outcomes were due to the intervention(s). | 2-7 |
| 10. **Measures** |  |
| a. Measures chosen for studying processes and outcomes of the intervention(s), including rationale for choosing them, their operational definitions and their validity and reliability. | 2-7 |
| b. Description of the approach to the ongoing assessment of contextual elements that contributed to the success, failure, efficiency and cost. | 2-7 |
| c. Methods employed for assessing completeness and accuracy of data. | 2-7 |
| 11. **Analysis** | 7-10 |
| a. Qualitative and quantitative methods used to draw inferences from the data. | 7-10 |
| b. Methods for understanding variation within the data, including the effects of time as a variable. | 7-10 |
| 12. **Ethical considerations** - Ethical aspects of implementing and studying the intervention(s) and how they were addressed, including, but not limited to, formal ethics review and potential conflict(s) of interest. | 12 |
| **Results: What did you find?** |  |
| 13. **Results** |  |
| a. Initial steps of the intervention(s) and their evolution over time (eg, time-line diagram, flow chart or table), including modifications made to the intervention during the project. | 7-10 |
| b. Details of the process measures and outcomes. | 7-10 |
| c. Contextual elements that interacted with the intervention(s). | 7-10 |
| d. Observed associations between outcomes, interventions and relevant contextual elements. | 7-10 |
| e. Unintended consequences such as unexpected benefits, problems, failures or costs associated with the intervention(s). | 11 |
| f. Details about missing data. | 11 |
| **Discussion: What does it mean?** | 7-10 |
| 14. **Summary** | 7-10 |
| a. Key findings, including relevance to the rationale and specific aims. | 7-10 |
| b. Particular strengths of the project. | 7-10 |
| 15. **Interpretation** | 7-11 |
| a. Nature of the association between the intervention(s) and the outcomes. | 7-11 |
| b. Comparison of results with findings from other publications. | 7-11 |
| c. Impact of the project on people and systems. | 7-11 |
| d. Reasons for any differences between observed and anticipated outcomes, including the influence of context. | 7-11 |
| e. Costs and strategic trade-offs, including opportunity costs. | 7-11 |
| 16. **Limitations** | 10-11 |
| a. Limits to the generalisability of the work. | 10-11 |
| b. Factors that might have limited internal validity such as confounding, bias or imprecision in the design, methods, measurement or analysis. | 10-11 |
| c. Efforts made to minimise and adjust for limitations. | 10-11 |
| **Conclusions** | 11-12 |
| a. Usefulness of the work. | 11-12 |
| b. Sustainability. | 11-12 |
| c. Potential for spread to other contexts. | 11-12 |
| d. Implications for practice and for further study in the field. | 11-12 |
| e. Suggested next steps. | 11-12 |
| **Other information** | 12 |
| 18. **Funding** - Sources of funding that supported this work. Role, if any, of the funding organisation in the design, implementation, interpretation and reporting. | 12 |
